# Biomarkers of bleeding and venous thromboembolism in patients with acute leukemia

**DOI:** 10.1101/2023.10.18.23297216

**Authors:** Yohei Hisada, Sierra J. Archibald, Karan Bansal, Yanjun Chen, Chen Dai, Sindhu Dwarampudi, Nora Balas, Lindsey Hageman, Nigel S. Key, Smita Bhatia, Ravi Bhatia, Nigel Mackman, Radhika Gangaraju

**Affiliations:** UNC Blood Research Center, Division of Hematology, Department of Medicine, University of North Carolina at Chapel Hill, Chapel Hill, USA; Institute for Cancer Outcomes and Survivorship, School of Medicine, University of Alabama at Birmingham, Birmingham, USA; Department of Medicine, School of Medicine, University of Alabama at Birmingham, Birmingham, USA

**Author notes:** Corresponding Authors: Yohei Hisada, Ph.D. UNC Blood Research Center, Department of Medicine University of North Carolina at Chapel Hill, 8004 Mary Ellen Jones Bldg. CB#7035,116 Manning Drive, Chapel Hill, NC 27599; Radhika Gangaraju, MD, MSPH, Institute for Cancer Outcomes and Survivorship, University of Alabama at Birmingham 1600 7^th^ Avenue South, Lowder 500, Birmingham, AL 35233.

**Keywords:** acute leukemia, biomarker, bleeding, disseminated intravascular coagulation, venous thrombosis

## Abstract

**Background:** Coagulopathy and associated bleeding and venous thromboembolism (VTE) are major causes of morbidity and mortality in patients with acute leukemia. The underlying mechanisms of these complications have not been fully elucidated.

**Objectives:** To evaluate the associations between biomarker levels and bleeding and VTE in acute leukemia patients.

**Patients/Method:** We examined plasma levels of activators, inhibitors and biomarkers of the coagulation and fibrinolytic pathways in patients ≥18 years with newly diagnosed acute leukemia compared to healthy controls. Multivariable regression models were used to examine the association of biomarkers with bleeding and VTE in acute leukemia patients. The study included 358 patients with acute leukemia (29 acute promyelocytic leukemia [APL], 253 non-APL acute myeloid leukemia [AML] and 76 acute lymphoblastic leukemia [ALL]), and 30 healthy controls.

**Results:** Patients with acute leukemia had higher levels of extracellular vesicle (EV) tissue factor (TF) activity, phosphatidylserine-positive EVs, plasminogen activator inhibitor-1 (PAI-1), plasmin-antiplasmin complexes, cell-free DNA and lower levels of citrullinated histone H3-DNA complexes compared to healthy controls. APL patients had the highest levels of EVTF activity and the lowest levels of tissue plasminogen activator among the acute leukemia patients. There were 41 bleeding and 37 VTE events in acute leukemia patients. High EVTF activity was associated with increased risk of bleeding (sHR 2.30, 95%CI 0.99-5.31) whereas high PAI-1 was associated with increased risk of VTE (sHR 3.79, 95%CI 1.40-10.28) in these patients.

**Conclusions:** Our study shows alterations in several biomarkers in acute leukemia and identifies biomarkers associated with risk of bleeding and VTE.

**Essentials:**

1. The mechanisms of acute leukemia-associated bleeding and thrombosis have not been elucidated.
2. We measured plasma biomarkers of coagulation and fibrinolysis in acute leukemia patients.
3. Biomarkers of the coagulation and fibrinolytic pathways are altered in acute leukemia patients.
4. EVTF activity is associated with bleeding, and PAI-1 is associated with VTE in acute leukemia.

## Introduction

Acute leukemia is a rapidly progressing hematologic malignancy and includes two distinct major morphologic types, acute myeloid leukemia (AML) and acute lymphoblastic leukemia (ALL). While the 5-year survival rates are improving for acute leukemia, early morbidity and mortality from disseminated intravascular coagulation (DIC) and associated complications, such as bleeding and venous thromboembolism (VTE) remain high.[1–12] AML and ALL have a similar incidence of DIC at 9-22%.[13–18] Acute promyelocytic leukemia (APL), a subtype of AML, has a higher incidence of DIC (60-85%)[19–21] with high incidence of early mortality (7-26% within 30 days of leukemia diagnosis)[4–8, 22–26] compared to other types of acute leukemia. A recent study found that high white blood cell (WBC) count is associated with early hemorrhagic death in APL patients and indicated a role of leukemia cells in coagulopathy.[27]

Tissue factor (TF) is a transmembrane receptor for factor (F) VII/VIIa. The TF/FVIIa complex is the major physiological initiator of the coagulation protease cascade.[28] Leukemia cells express TF.[29–32] Peripheral blood mononuclear cells (PBMCs that included leukemic blasts) from AML patients were found to express higher TF procoagulant activity compared with PBMCs from healthy controls;[29] notably TF procoagulant activity was higher in APL patients compared to non-APL AML patients.[29] In addition, PBMCs from AML patients with DIC had higher levels of TF activity compared to those from patients without DIC.[29] A human APL cell line NB4 and a human AML cell line HL-60 express TF, with higher levels in NB4 cells.[30–32] All-trans retinoic acid (ATRA) reduced TF expression in bone marrow cells (including blast cells) from APL patients and NB4 cells.[30, 33–37]

Many cells, including cancer cells and activated monocytes, release submicron vesicles called extracellular vesicles (EVs) that express TF and expose procoagulant phospholipids, such as phosphatidylserine (PS), on their surface.[38, 39] NB4 and HL-60 cells have been shown to release TF+EVs.[32] Studies with small numbers of patients have shown high levels of circulating EVTF activity in AML patients with DIC.[40–42] APL is also associated with hyperfibrinolysis.[43] APL patients have higher levels of urokinase-type plasminogen activator receptor (uPAR) and plasmin-antiplasmin complexes (PAP) compared to healthy controls.[44] In contrast, levels of plasminogen activator inhibitor-1 (PAI-1) antigen were lower in APL patients compared with controls.[44] Another study found that PAI-1 activity was lower in APL patients with DIC compared to healthy controls.[45] However, these studies were limited by small sample sizes (1-24 patients). To date, there are no studies that have examined the association between biomarkers in coagulation and fibrinolytic pathways with bleeding and VTE using a large number of samples of acute leukemia patients.

Leukemic blasts release cell-free (cf) DNA into the circulation.[46] Indeed, patients with acute leukemia have higher levels of cfDNA compared to healthy population.[29, 47] cfDNA can activate FXII, which leads to activation of the coagulation system.[48] Activated neutrophils release neutrophil extracellular traps (NETs) that consist of extracellular chromatin components, including DNA, citrullinated histone H3 (H3Cit) and neutrophil granule proteins.[49] NETs enhance VTE by capturing platelets and procoagulant EVs.[50–53] However, recent studies reported that neutrophils and granulocytes isolated from acute leukemia patients had reduced capacity to form NETs compared to those from healthy controls.[54–56]

The current study had two goals. The first was to compare levels of activators, inhibitors and biomarkers of coagulation and fibrinolysis in patients with acute leukemia with healthy controls. The second was to determine the association between these biomarkers with bleeding and VTE in acute leukemia patients.

## Methods

### Study Population

This prospective cohort study included adult patients (≥18 years at diagnosis) with newly diagnosed acute leukemia who provided blood samples for the University of Alabama at Birmingham (UAB) biorepository between May 2016 and April 2021. Of the 826 subjects enrolled in the study, 570 had a diagnosis of acute leukemia. Of these, patients with relapsed/refractory disease at the time of sample collection, insufficient clinical data, and those who did not have adequate plasma sample were excluded, yielding a final sample size of 358 (**Supplementary Figure 1**). Blood samples were collected in 10 mL EDTA tubes on the day of diagnosis and centrifuged at 2000 rpm for 5 minutes. The plasma was separated and stored at - 80°C. Details of patients’ characteristics were described in the supplementary materials.

### Patients’ characteristics

Demographic information, clinical data regarding leukemia diagnosis, comorbidities, chemotherapy, and outcomes (VTE and bleeding) were abstracted from the medical records until blood or marrow transplantation, last contact with patient or death, whichever occurred first. Laboratory values as part of routine clinical care included WBC count, hemoglobin, platelet count, prothrombin time (PT), partial thromboplastin time (PTT), D-dimer, fibrinogen, creatinine, lactate dehydrogenase (LDH) and peripheral blood blast cell percentage at leukemia diagnosis. This study was conducted with the approval of the institutional review board at UAB and informed consent was obtained according to the Declaration of Helsinki.

### Healthy controls

Plasma samples were obtained from 30 healthy individuals from whole blood drawn into EDTA tubes from Innovative Research (Cat# IPLASK2E2ML, Novi, MI, USA) using the same methodology as leukemia patients.

### Biomarkers

The transfer to the University of North Carolina at Chapel Hill and measurement of biomarkers was approved by the Institutional Review Board of the University of North Carolina at Chapel Hill (21-0554). Biomarkers in the coagulation and fibrinolytic pathways, and NETs were measured at the University of North Carolina at Chapel Hill. EVTF activity were measured using an in-house 2-stage FXa generation assay as described previously.[57] PS+ EVs were measured using the ZYMUPHEN^TM^ MP-ACTIVITY assay kit (Cat# 521096, Aniara Diagnostica, West Chester, OH, USA). H3Cit-DNA complexes were measured using an in-house ELISA as described previously.[58] cfDNA were measured using the Quant-iT^TM^ PicoGreen^TM^ dsDNA Assay Kit (Cat#P11496, Thermo Fisher Scientific, Waltham, MA, USA). PAP (Cat#MBS2503292, MyBioSource, San Diego, CA, USA), tissue plasminogen activator (tPA, Cat#IHUTPAKTT, Innovative Research, Novi, MI, USA) and active PAI-1 (Cat#HPAIKT, Innovative Research) were measured using commercial assays. EVTF activity data from 6 patients were used in a publication[59] for comparing our in-house EVTF activity assay and commercial TF ELISAs.

### Outcomes and characteristics

Outcomes and characteristics of interest for this study included overt DIC at the time of acute leukemia diagnosis, bleeding and VTE. Overt DIC was defined based on the scoring system developed by the ISTH, using levels of D-dimer, fibrinogen, PT, and platelet count at the time of initial diagnosis.[60, 61] Bleeding episodes were defined as major and clinically relevant non-major bleeding as per ISTH criteria.[62] VTE events included radiologically confirmed symptomatic or incidental pulmonary embolism, proximal or distal lower extremity deep vein thrombosis (DVT), upper extremity DVT, DVT at other sites, and superficial venous thrombosis including catheter-related thrombosis.[63] Arterial thrombosis events were excluded. Detailed definitions of outcomes are provided in **Supplementary Table 1**. A trained reviewer (KB) performed abstraction of all radiology reports, clinical notes, and discharge summaries and each outcome was independently assessed by a second reviewer (SD). Discrepancies were resolved by a third clinician reviewer with expertise in hematology (RG). Outcomes were evaluated from the index date of acute leukemia diagnosis until blood or bone marrow transplantation, last follow-up or death.

### Statistical Analyses

The Shapiro-Wilk test was used for normality testing. For biomarker comparisons between leukemia patients and controls, we used the unpaired 2-tailed Student *t* test or the Mann-Whitney U test depending on normality. The Kruskal-Wallis test with the Dunn’s multiple comparisons test was used to compare biomarker levels in multiple groups. These analyses were performed with PRISM version 7.03 (GraphPad Software, La Jolla, CA, USA). We used SAS software v9.4 (SAS Institute Inc., Cary, North Carolina, USA) to analyze biomarker associations with bleeding and VTE outcomes. Age at leukemia diagnosis and body mass index (BMI) were treated as continuous variables; the remaining variables were considered categorical. We calculated cumulative incidence of bleeding or VTE, using Fine and Gray methods, treating death as a competing risk.[64] We used two-sample *t* test or Wilcoxon rank-sum test (for continuous variables) and chi-square test (for categorical variables) to compare differences between groups. Using proportional sub-distribution hazards multivariable regression analysis,[65] we examined the association between biomarkers and the outcomes of interest. For this analysis, biomarkers were considered as tertiles and third and second tertile distributions were compared to the first tertile, and adjusted for clinically relevant covariates in multivariable models. Due to collinearity between biomarkers, we developed multiple individual multivariable models for each biomarker adjusting for the relevant covariates. For bleeding outcome, we created two multivariable models for each biomarker; model 1 adjusted for age at leukemia diagnosis, sex and race/ethnicity and model 2 for age, sex, race/ethnicity, ISTH DIC score, comorbidities (hypertension, diabetes, dyslipidemia, congestive heart failure, valvular heart disease, rheumatologic disorder, peripheral vascular disease, peptic ulcer disease, inflammatory bowel disease, chronic obstructive pulmonary disease, arrhythmia, psychiatric disorder, liver disease, human immunodeficiency virus [HIV] infection, chronic kidney disease, history of prior transplant, coronary heart disease, stroke) and history of bleeding. For VTE risk, multivariable models were adjusted for age at leukemia diagnosis, sex, race/ethnicity, acute leukemia type, BMI, history of VTE, and comorbidities We also performed sub-analyses restricted to AML patients. Two-sided tests with *p* <0.05 were considered statistically significant. Statistical analyses are described in the supplementary materials.

### Data sharing statement

For original data, please contact

## Results

### Patient characteristics

Demographic and clinical characteristics of acute leukemia patients and controls as well as routine laboratory parameters at leukemia diagnosis are shown in **Table 1**. The study included 358 patients with acute leukemia (29 APL, 253 non-APL AML, 76 ALL) and 30 controls. The median age at diagnosis of acute leukemia was 59 years (range: 19-89). The controls were enrolled at a median age of 58.5 years (range: 23-76). There were 203 (56.7%) males and 264 (73.7%) non-Hispanic white individuals with acute leukemia. Over a median follow up of 11.9 months (range: 0.12-78.1 months), 41 acute leukemia patients developed bleeding (23 with major bleeding, 18 with clinically relevant non-major bleeding) and 37 developed VTE (24 DVT and 13 patients with superficial vein thromboses). It is notable that 28 (75.7%) of these VTE events were catheter related. There were 7 APL (24.1%), 31 non-APL AML (12.3%) and 3 ALL (3.9%) patients who developed bleeding and 3 APL (10.3%), 30 non-APL AML (11.9%) and 4 ALL (5.3%) patients who developed VTE. Characteristics of bleeding and VTE events are shown in **Table 2**.

**Table 1.**
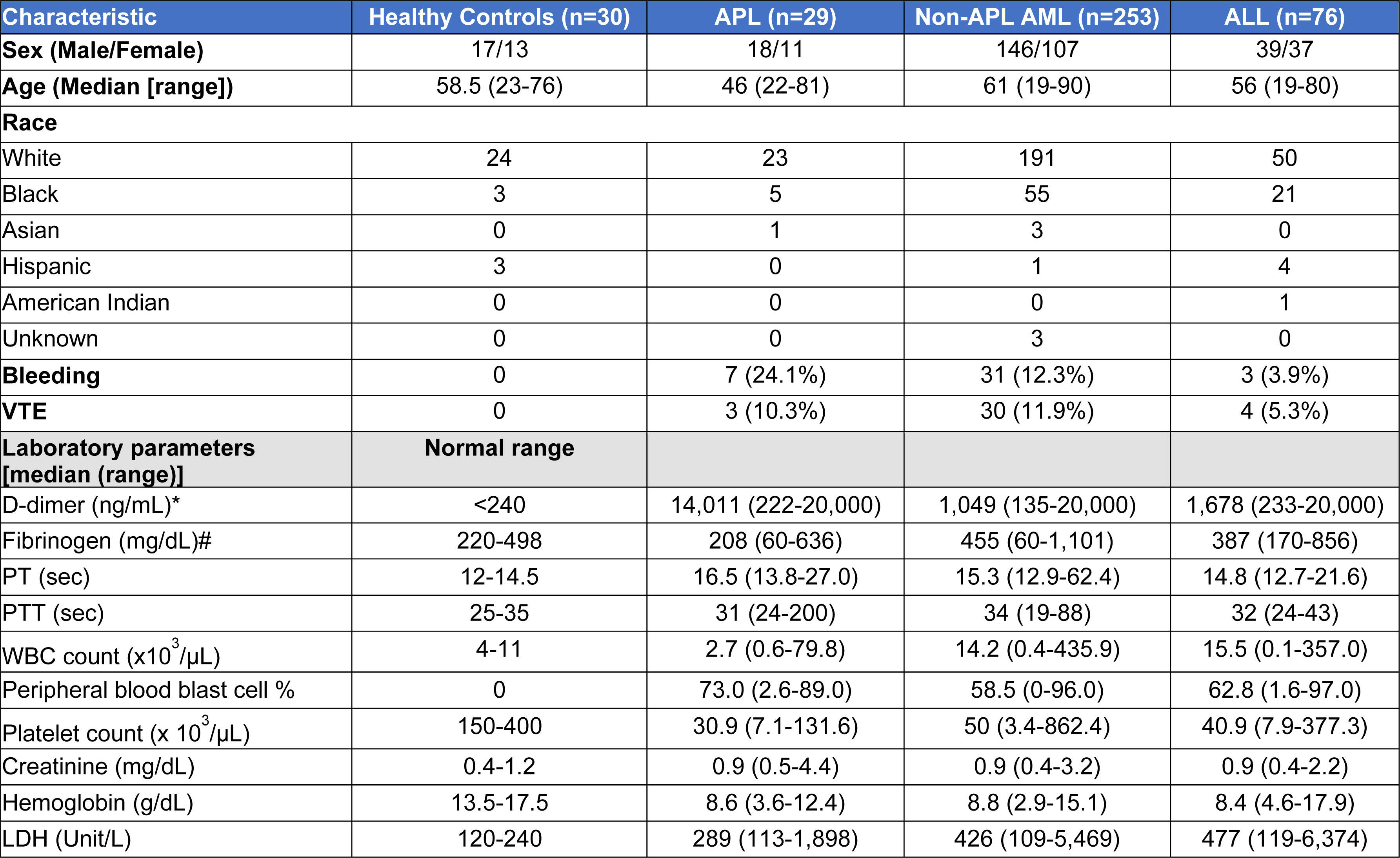
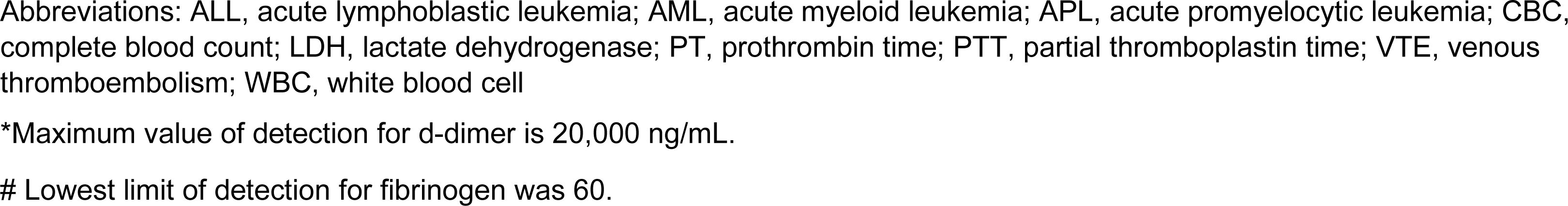
Demographic characteristics of healthy controls and acute leukemia patients and laboratory values for acute leukemia patients at diagnosis.

| Characteristic | Healthy Controls (n=30) | APL (n=29) | Non-APL AML (n=253) | ALL (n=76) |
| --- | --- | --- | --- | --- |
| **Sex (Male/Female)** | 17/13 | 18/11 | 146/107 | 39/37 |
| **Age (Median [range])** | 58.5 (23-76) | 46 (22-81) | 61 (19-90) | 56 (19-80) |
| **Race** |  |  |  |  |
| White | 24 | 23 | 191 | 50 |
| Black | 3 | 5 | 55 | 21 |
| Asian | 0 | 1 | 3 | 0 |
| Hispanic | 3 | 0 | 1 | 4 |
| American Indian | 0 | 0 | 0 | 1 |
| Unknown | 0 | 0 | 3 | 0 |
| **Bleeding** | 0 | 7 (24.1%) | 31 (12.3%) | 3 (3.9%) |
| **VTE** | 0 | 3 (10.3%) | 30 (11.9%) | 4 (5.3%) |
| **Laboratory parameters**<br>**[median (range)]** | **Normal range** |  |  |  |
| D-dimer (ng/mL)* | <240 | 14,011 (222-20,000) | 1,049 (135-20,000) | 1,678 (233-20,000) |
| Fibrinogen (mg/dL)# | 220-498 | 208 (60-636) | 455 (60-1,101) | 387 (170-856) |
| PT (sec) | 12-14.5 | 16.5 (13.8-27.0) | 15.3 (12.9-62.4) | 14.8 (12.7-21.6) |
| PTT (sec) | 25-35 | 31 (24-200) | 34 (19-88) | 32 (24-43) |
| WBC count ($\times 10^3/\mu\text{L}$) | 4-11 | 2.7 (0.6-79.8) | 14.2 (0.4-435.9) | 15.5 (0.1-357.0) |
| Peripheral blood blast cell % | 0 | 73.0 (2.6-89.0) | 58.5 (0-96.0) | 62.8 (1.6-97.0) |
| Platelet count ($\times 10^3/\mu\text{L}$) | 150-400 | 30.9 (7.1-131.6) | 50 (3.4-862.4) | 40.9 (7.9-377.3) |
| Creatinine (mg/dL) | 0.4-1.2 | 0.9 (0.5-4.4) | 0.9 (0.4-3.2) | 0.9 (0.4-2.2) |
| Hemoglobin (g/dL) | 13.5-17.5 | 8.6 (3.6-12.4) | 8.8 (2.9-15.1) | 8.4 (4.6-17.9) |
| LDH (Unit/L) | 120-240 | 289 (113-1,898) | 426 (109-5,469) | 477 (119-6,374) |

Abbreviations: ALL, acute lymphoblastic leukemia; AML, acute myeloid leukemia; APL, acute promyelocytic leukemia; CBC, complete blood count; LDH, lactate dehydrogenase; PT, prothrombin time; PTT, partial thromboplastin time; VTE, venous thromboembolism; WBC, white blood cell
\*Maximum value of detection for d-dimer is 20,000 ng/mL.

**Table 2.**
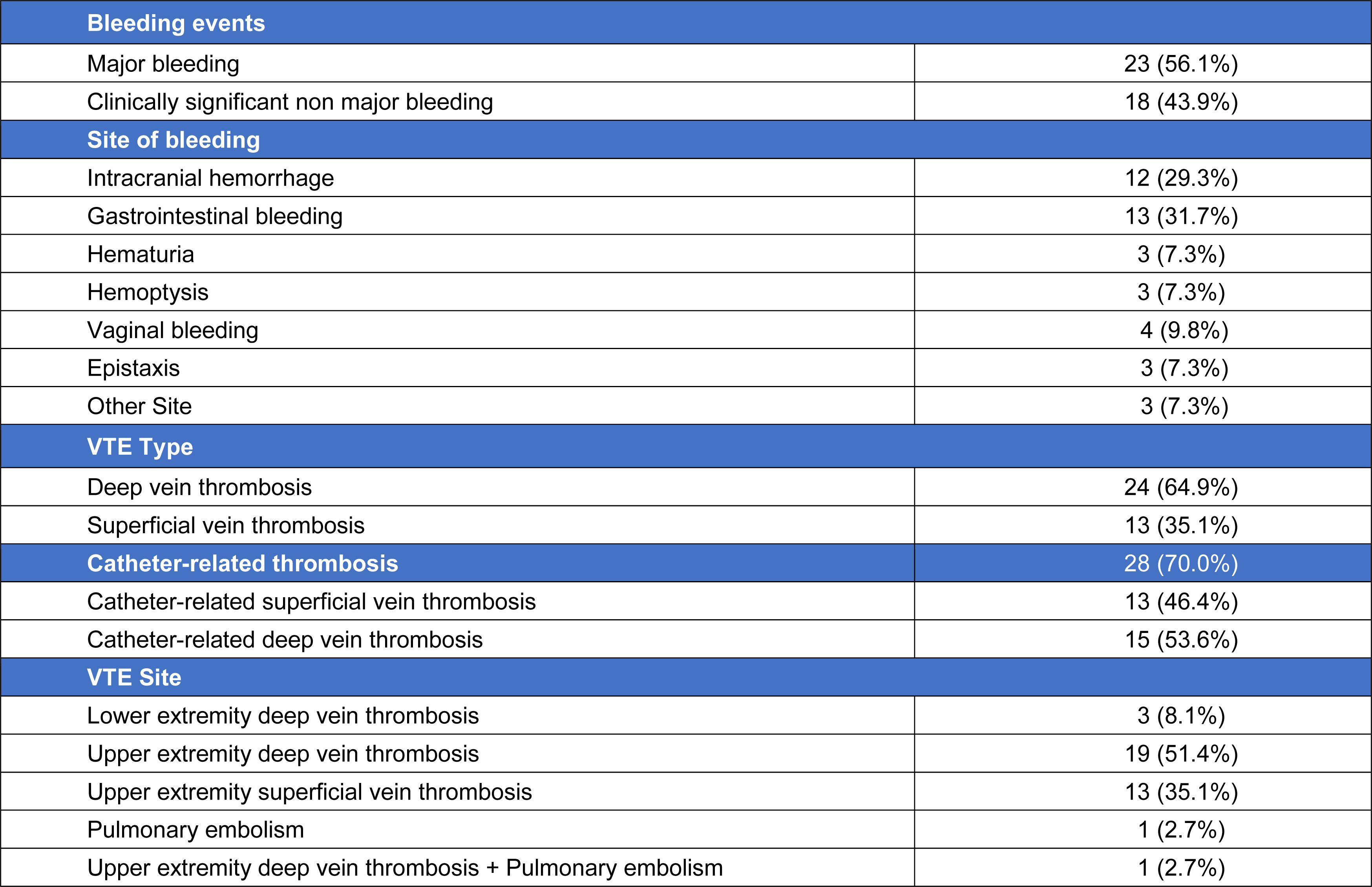
Characteristics of bleeding and venous thromboembolism events.

### Biomarker levels in acute leukemia patients compared to controls

#### Procoagulant EVs, cfDNA and a biomarker of neutrophil extracellular traps

EVTF activity was higher in leukemia patients compared to healthy controls (**Figure 1A**). APL patients had the highest levels of EVTF activity compared to non-APL AML or ALL patients. Higher levels of PS+EVs were observed in all three leukemia types compared to healthy controls; no significant differences were observed among leukemia type (**Figure 1B**). Levels of cfDNA were higher in leukemia patients compared to healthy controls (**Figure 1C**), with the highest levels present in those with ALL. Interestingly, H3Cit-DNA complexes levels were significantly lower in all three leukemia types compared to healthy controls (**Figure 1D**).

**Figure 1.**
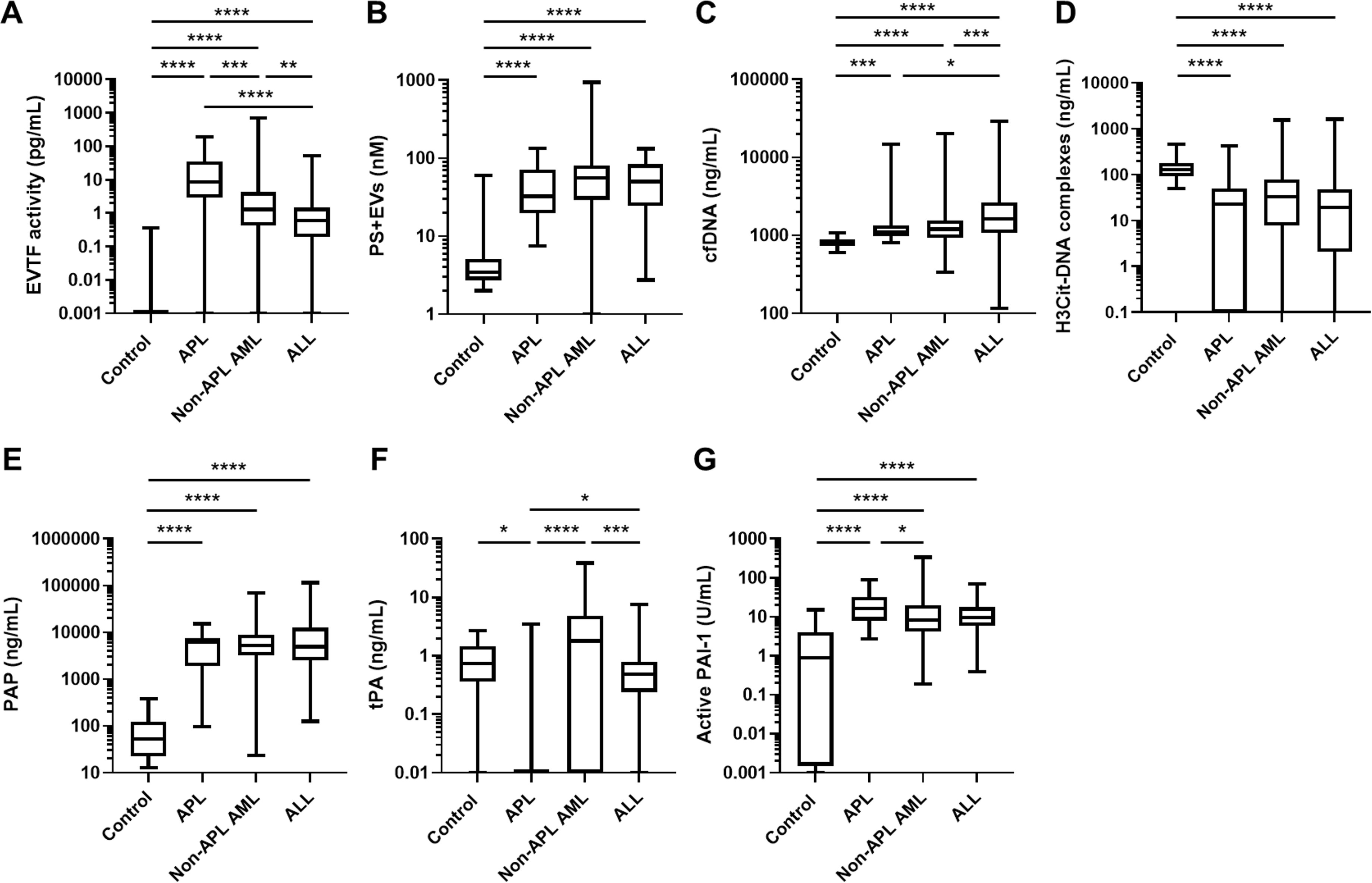
Levels of biomarkers in the coagulation pathway, neutrophil extracellular trap formation and the fibrinolytic pathway in patients with different types of acute leukemia. Levels of (A) extracellular vesicle (EV) tissue factor (TF) activity, (B) phosphatidylserine-positive (PS+) EVs, (C) cell-free (cf) DNA and (D) citrullinated histone H3 (H3Cit)-DNA complexes (E) plasmin-antiplasmin complex(PAP), (F) tissue plasminogen activator (tPA) and (G) plasminogen activator inhibitor-1 (PAI-1) in patients with acute promyelocytic leukemia (APL), non-APL acute myeloid leukemia (AML), and acute lymphoblastic leukemia (ALL) were measured. Data are shown as box and whisker plots. The box ranges from the first quartile to the third quartile of the distribution. The median is indicated by a line across the box. The whiskers on box plots extend to the minimum and maximum data points. \**P* < 0.05, \*\**P* < 0.01, \*\*\**P* < 0.001, \*\*\*\**P* < 0.0001.

#### Biomarkers of the fibrinolytic pathway

Levels of PAP were significantly higher in the leukemia patients compared to healthy controls (**Figure 1E**). tPA levels were significantly lower in APL patients compared to healthy controls, as well as non-APL AML and ALL patients (**Figure 1F**). PAI-1 levels were higher in leukemia patients compared to controls with significantly higher levels in the APL group compared to the non-APL AML group (**Figure 1G**).

### Comparisons between acute leukemia patients with high and mid to low EVTF activity

Next, we determined if EVTF activity is associated with leukocytosis, high percentage of blast cells, D-dimer and platelet count in acute leukemia patients. We compared leukocyte count, peripheral blood blast cell percentage, D-dimer levels and platelet count between acute leukemia patients with the top 10% of EVTF activity (referred to as high EVTF, n = 36) and the rest of acute leukemia patients (referred to as mid to low EVTF, n = 322). We found that both WBC count and peripheral blood blast cell percentage were significantly higher in the high EVTF activity group compared to the mid to low EVTF activity group (**Supplementary Table 2**). We also found that D-dimer levels were significantly higher in the high EVTF activity group compared to the mid to low EVTF activity group (**Supplementary Table 2**). In contrast, we did not see a difference in platelet counts between the high EVTF activity group and the mid to low EVTF activity group (**Supplementary Table 2**).

### Comparisons between leukemia patients with and without overt DIC

We calculated the ISTH overt DIC score and found that 90% of APL patients, 34% of non-APL AML patients, and 45% of ALL patients had overt DIC at diagnosis. EVTF activity and WBC count were significantly higher in patients with APL with overt DIC compared to patients without DIC (referred to as no-DIC) (**Supplementary Table 3**). In patients with APL, the ISTH overt DIC score components (D-dimer, fibrinogen, PT, and platelets) were significantly different between with and without overt DIC patients. In patients with non-APL AML, we found that EVTF activity, cfDNA, PTT, WBC count, peripheral blood blast cell percentage, creatinine, and LDH were significantly higher in patients with overt DIC compared to patients without DIC (**Supplementary Table 4**). In patients with non-APL AML, the three ISTH overt DIC score components (D-dimer, PT, and platelets) were significantly different between patients with and without overt DIC. The incidence of bleeding was significantly higher in non-APL AML patients with overt DIC compared to non-APL AML patients without DIC patients. In patients with ALL, we found significantly higher levels of PAP and lower levels of hemoglobin in patients with overt DIC compared to patients without DIC. PS+EVs levels were lower in ALL patients with overt DIC compared to patients without DIC (**Supplementary Table 5**). D-dimer and platelets were the only components of the ISTH overt DIC score that showed significant differences between ALL patients with overt DIC and those without DIC.

### Bleeding risk and associated biomarkers in acute leukemia patients

The characteristics of acute leukemia patients with and without bleeding are shown in **Supplementary Table 6**. Patients with bleeding had higher D-dimer and LDH levels, and prolonged PT compared to those without. The cumulative incidence of bleeding was 6.0% at 1 month (standard deviation [SD]: 1.3) and 12.6% at 1 year (SD: 1.9) (**Figure 2A**). The majority of the bleeding events in APL patients occurred within 1 month after diagnosis with a cumulative incidence of 25.0±8.4% at 1 month, whereas the cumulative incidence was 5.3±1.4% at 1 month and 14.6±2.6% at 1 year in non-APL AML patients and 1.4±1.3% at 1 month and 2.9±2.1% at 1 year in ALL patients (**Figure 2B**).

**Figure 2.**
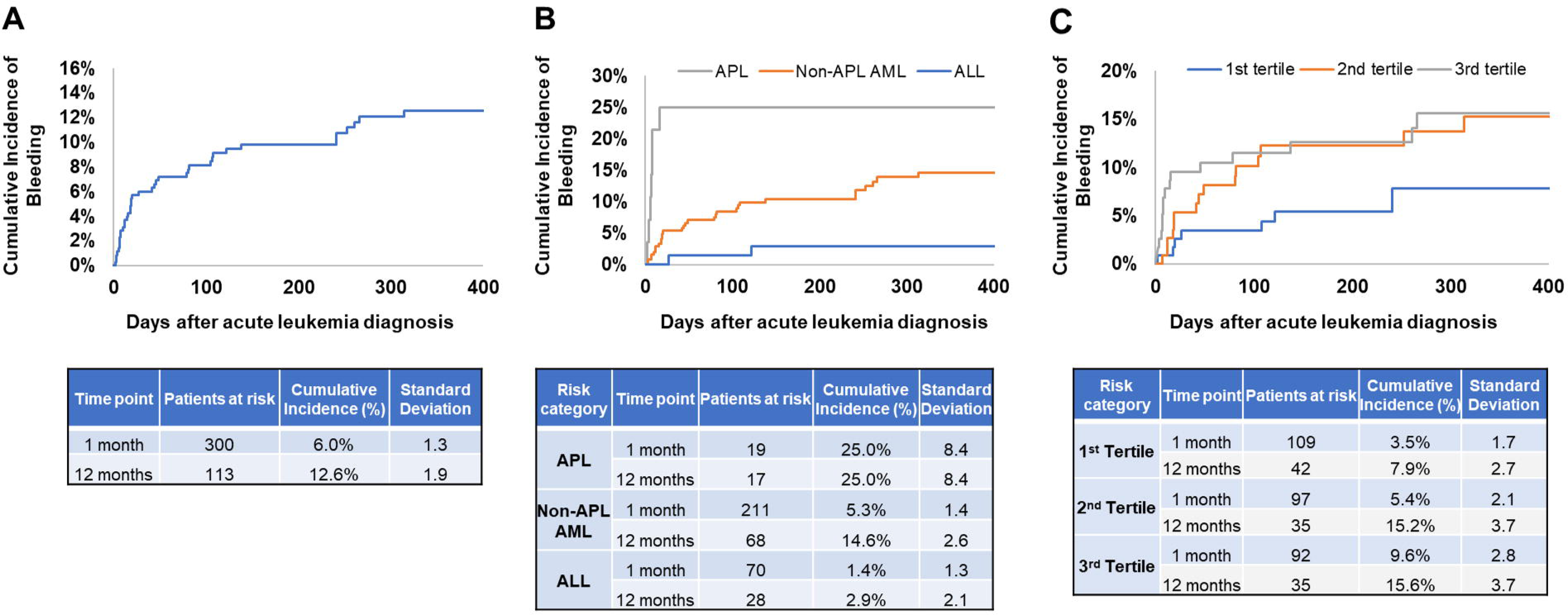
Cumulative incidence of bleeding in patients with different types of acute leukemia. (A) Data from all patients, (B) Cumulative incidence by acute leukemia type and (C) Cumulative incidence by tertiles of extracellular vesicle tissue factor activity were shown.

When examining the associations between biomarkers and bleeding, we combined all leukemia types to ensure sufficient statistical power. Multivariable sub-distribution hazards model with death as competing risk and adjusting for age, sex, race/ethnicity showed that EVTF activity (sHR 2.33, 95%CI 1.08-5.04), PT (sHR 2.55, 95%CI 1.15-5.62), WBC count (sHR 2.30, 95%CI 1.03-5.13), and LDH (sHR 2.80, 95%CI 1.12-6.97) were associated with increased bleeding risk (**Table 3**). After further adjustment for the ISTH DIC score, comorbidities and history of bleeding, only EVTF activity (sHR 2.30, 95%CI 0.99-5.31) was associated with increased risk of bleeding. The cumulative incidence of bleeding at 1 year was 15.6±3.7%, 15.2±3.7% and 7.9±2.7% for patients with EVTF activity in the third, second and first tertiles, respectively (**Figure 2C**). In addition, we set the median level of each biomarker as a cut-off value and calculated the percentage of above median in acute leukemia patients with and without bleeding. Acute leukemia patients with bleeding had a higher percentage of above median levels of PT (without vs with bleeding: 48.9 vs 56.1%), WBC (without vs with bleeding: 47.6 vs 68.3%), LDH (47.5 vs 66.7%) and EVTF activity (49.0 vs 56.1%) compared to acute leukemia patients without bleeding (**Supplementary Figure 2 A-D**).

**Table 3.** Biomarkers associated with bleeding risk in acute leukemia.

| Biomarker | Univariable analysis |  | Multivariable analysis |  |  |  |
| --- | --- | --- | --- | --- | --- | --- |
|  | HR (95% CI) | P value | Model 1* |  | Model 2# |  |
|  |  |  | HR (95% CI) | P value | HR (95% CI) | P value |
| EVTF activity | **2.32 (1.08 - 4.99)** | **0.032** | **2.33 (1.08 - 5.04)** | **0.031** | **2.30 (0.99 - 5.31)** | **0.052** |
| PS + EVs | 1.11 (0.57 - 2.16) | 0.755 | 1.11 (0.57 - 2.17) | 0.753 | 1.20 (0.58 - 2.49) | 0.627 |
| cfDNA | 1.62 (0.80 - 3.28) | 0.181 | 1.64 (0.83 - 3.26) | 0.153 | 1.80 (0.83 - 3.88) | 0.135 |
| H3Cit-DNA complexes | 0.85 (0.45 - 1.61) | 0.622 | 0.88 (0.46 - 1.68) | 0.703 | 0.94 (0.47 - 1.88) | 0.855 |
| PAP | 1.55 (0.76 - 3.18) | 0.226 | 1.46 (0.71 - 3.00) | 0.301 | 1.41 (0.66 - 3.02) | 0.376 |
| tPA | 0.84 (0.44 - 1.59) | 0.590 | 0.78 (0.40 - 1.53) | 0.476 | 0.82 (0.40 - 1.68) | 0.587 |
| PAI-1 | 1.21 (0.62 - 2.36) | 0.578 | 1.18 (0.59 - 2.36) | 0.646 | 1.15 (0.57 - 2.35) | 0.691 |
| D-dimer | 1.29 (0.64 - 2.57) | 0.476 | 1.21 (0.60 - 2.47) | 0.597 | NA | NA |
| Fibrinogen | 0.75 (0.40 - 1.41) | 0.365 | 0.72 (0.38 - 1.38) | 0.326 | NA | NA |
| PT | **2.54 (1.14 - 5.68)** | **0.023** | **2.55 (1.15 - 5.62)** | **0.021** | NA | NA |
| PTT | 0.87 (0.45 - 1.66) | 0.669 | 0.82 (0.42 - 1.58) | 0.547 | 0.83 (0.41 - 1.65) | 0.592 |
| WBC count | **2.17 (0.99 - 4.71)** | **0.052** | **2.30 (1.03 - 5.13)** | **0.041** | 2.30 (0.96 - 5.53) | 0.061 |
| Peripheral blood blast cell | 1.91 (0.88 - 4.15) | 0.104 | 1.96 (0.90 - 4.28) | 0.090 | 2.20 (0.89 - 5.40) | 0.086 |
| Platelet count | 0.56 (0.31 - 1.04) | 0.066 | **0.55 (0.30 - 1.02)** | **0.059** | NA | NA |
| Creatinine | 1.18 (0.59 - 2.38) | 0.642 | 1.25 (0.53 - 2.96) | 0.605 | 1.48 (0.57 - 3.85) | 0.422 |
| Hemoglobin | 0.65 (0.35 - 1.21) | 0.176 | 0.68 (0.37 - 1.25) | 0.215 | 0.64 (0.35 - 1.18) | 0.155 |
| LDH | **2.89 (1.21 - 6.90)** | **0.017** | **2.80 (1.12 - 6.97)** | **0.027** | 2.28 (0.92 - 5.63) | 0.075 |

\*Model 1 was adjusted for age, sex, race/ethnicity.
#Model 2 was adjusted for age, sex, race/ethnicity, ISTH DIC score (<5 vs ≥5), comorbidities (0, 1, ≥2), history of bleeding.
NA: not applicable as these labs are components of DIC score.
Abbreviations: cfDNA, cell-free DNA; EVTF, extracellular vesicle tissue factor; H3Cit-DNA, citrullinated histone H3-DNA; LDH, lactate dehydrogenase; PAI-1, plasminogen activator inhibitor-1; PAP, plasmin-antiplasmin complex; PS+EVs, phosphatidylserine-positive extracellular vesicles; PT, prothrombin time; PTT, partial thromboplastin time; tPA, tissue plasminogen activator; WBC, white blood cell; 95% CI, 95% confidence interval

### VTE risk and associated biomarkers in acute leukemia patients

The characteristics of acute leukemia patients with and without VTE are shown in **Supplement Table 7**. Patients with VTE had higher PAI-1 levels compared to those without. The cumulative incidence of overall VTE was 6.5±1.3% at 1 month and 11.3±1.8% at 1 year (**Figure 3A**). The cumulative incidence of VTE at 1 year was 10.4±5.8% in APL, 12.9±2.2% in non-APL AML and 7.3±3.8% in ALL (**Figure 3B**). High PAI-1 levels were associated with increased risk of VTE (sHR 3.79, 95%CI 1.40-10.28) (**Table 4**), and DVT (sHR 3.00, 95%CI 0.95-9.47) in multivariable models adjusting for age at leukemia diagnosis, sex, race/ethnicity, acute leukemia type, BMI, history of VTE and comorbidities. The cumulative incidence of VTE at 1 year was 17.5±3.7% in patients with PAI-1 levels in the third tertile of distribution compared to 12.2±3.3% in the second tertile and 4.4±1.9% in the first tertile (**Figure 3C**). The risk of DVT was also elevated in patients with elevated PAI-1 levels, with cumulative incidence of 12.3±3.3% in the third tertile vs 3.5±1.7% in the first tertile at 1 year (**Supplementary Figure 3**). High levels of cfDNA (sHR, 0.53; 95% CI, 0.28-1.00) and tPA (sHR, 0.45; 95% CI, 0.24-0.86) were associated with a decreased risk of VTE in univariable analysis but did not reach statistical significance in a multivariable model (**Table 4**). In addition, acute leukemia patients with VTE had a higher percentage of above median levels of PAI-1 compared to acute leukemia patients without VTE (without vs with bleeding: 47.6 vs 68.3%, **Supplementary Figure 4**).

**Figure 3.**
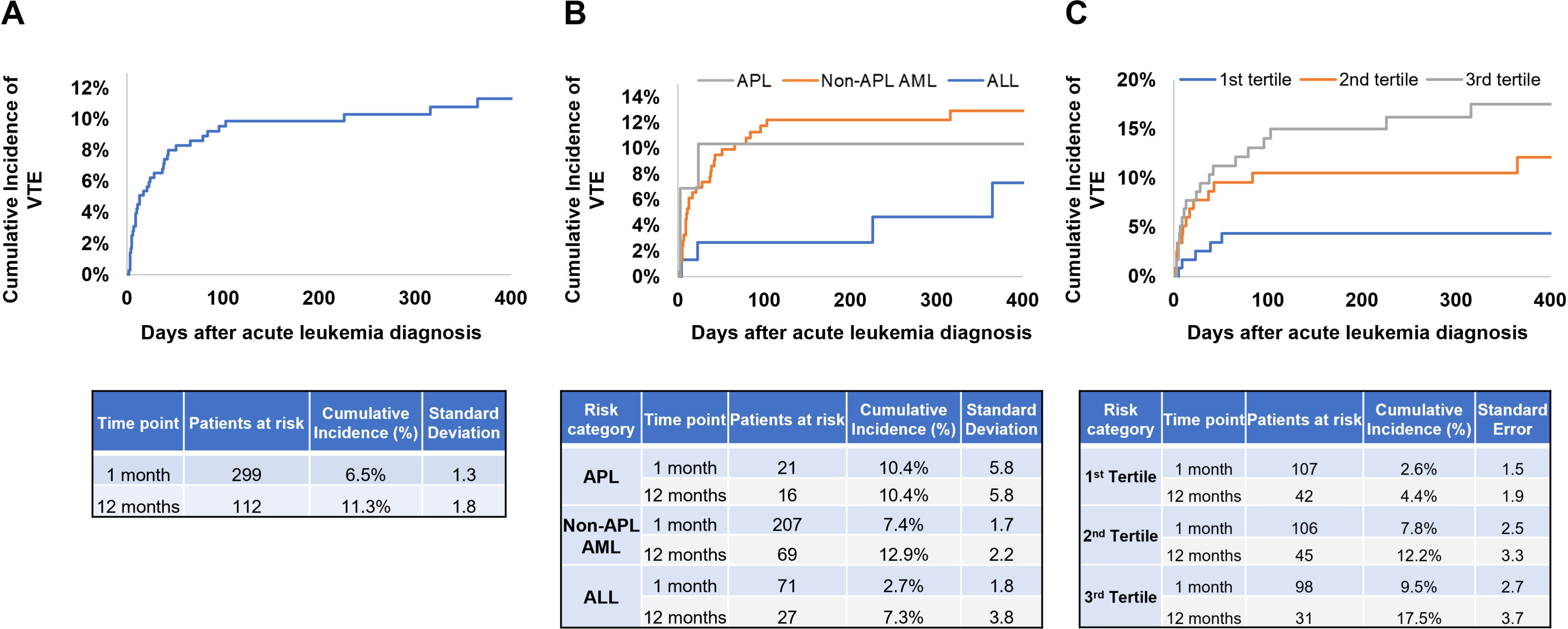
Cumulative incidence of venous thromboembolism in patients with different types of acute leukemia. (A) Data from all patients, (B) Cumulative incidence by acute leukemia type and (C) Cumulative incidence by tertiles of plasminogen activator inhibitor-1 levels were shown.

**Table 4.** Biomarkers associated with venous thromboembolism risk in acute leukemia.

| Biomarker | Univariable |  | Multivariable analysis* |  |
| --- | --- | --- | --- | --- |
|  | sHR (95% CI) | P value | sHR (95% CI) | P value |
| EVTF activity | 1.37 (0.67 - 2.81) | 0.395 | 1.28 (0.60 - 2.71) | 0.525 |
| PS + EVs | 0.84 (0.42 - 1.66) | 0.611 | 0.85 (0.42 - 1.72) | 0.648 |
| cfDNA | **0.53 (0.28 - 1.00)** | **0.051** | 0.62 (0.32 - 1.18) | 0.146 |
| H3Cit-DNA complexes | 0.71 (0.37 - 1.38) | 0.312 | 0.68 (0.34 - 1.36) | 0.277 |
| PAP | 0.82 (0.42 - 1.58) | 0.546 | 0.94 (0.45 - 1.95) | 0.863 |
| tPA | **0.45 (0.24 - 0.86)** | **0.015** | 0.51 (0.25 - 1.06) | 0.072 |
| PAI-1 | **3.36 (1.31 - 8.61)** | **0.011** | **3.79 (1.40 - 10.28)** | **0.009** |
| D-dimer | 0.79 (0.41 - 1.54) | 0.493 | 0.99 (0.45 - 2.17) | 0.980 |
| Fibrinogen | 0.85 (0.44 - 1.63) | 0.617 | 0.77 (0.38 - 1.57) | 0.472 |
| PT | 1.62 (0.77 - 3.42) | 0.207 | 1.53 (0.70 - 3.32) | 0.282 |
| PTT | 1.52 (0.72 - 3.22) | 0.271 | 1.52 (0.67 - 3.43) | 0.312 |
| WBC count | 1.06 (0.53 - 2.10) | 0.868 | 1.01 (0.49 - 2.05) | 0.986 |
| Peripheral blood blast cell | 1.11 (0.53 - 2.33) | 0.777 | 1.36 (0.64 - 2.92) | 0.425 |
| Platelet count | 0.93 (0.47 - 1.81) | 0.821 | 0.96 (0.48 - 1.92) | 0.913 |
| Creatinine | 1.23 (0.58 - 2.58) | 0.591 | 1.53 (0.67 - 3.50) | 0.309 |
| Hemoglobin | 1.43 (0.69 - 2.95) | 0.334 | 1.32 (0.65 - 2.70) | 0.446 |
| LDH | 0.91 (0.46 - 1.78) | 0.776 | 0.95 (0.48 - 1.87) | 0.882 |

\*Multivariable model was adjusted for age, sex, race/ethnicity, leukemia type, body mass index, history of venous thromboembolism and comorbidities.
Abbreviations: cfDNA, cell-free DNA; EVTF, extracellular vesicle tissue factor; H3Cit-DNA, citrullinated histone H3-DNA; LDH, lactate dehydrogenase; PAI-1, plasminogen activator inhibitor-1; PAP, plasmin-antiplasmin complex; PS+EVs, phosphatidylserine-positive extracellular vesicles; PT, prothrombin time; PTT, partial thromboplastin time; tPA, tissue plasminogen activator; WBC, white blood cell; 95% CI, 95% confidence interval

### Biomarkers of bleeding and VTE risk in non-APL AML patients

In a sub-analysis of patients with non-APL AML, EVTF activity (sHR 2.35; 95% CI, 0.98-5.66), WBC count (sHR 2.42; 95% CI, 1.04-5.62) and LDH (sHR 3.50; 95% CI, 1.34-9.17) were associated with bleeding risk. Low levels of tPA (sHR 0.47; 95% CI, 0.24-0.93) and high levels of PAI-1 (sHR 4.18; 95% CI, 1.47-11.87) were associated with VTE risk **(Supplementary Table 8)**.

### Biomarkers of intracranial hemorrhage in acute leukemia patients

Although the study was not designed to perform sub-analysis for intracranial hemorrhage and this sub-analysis is underpowered, we found that low levels of tPA and fibrinogen were associated with intracranial hemorrhage, and high EVTF activity was approaching statistical significance. (**Supplementary Table 9**).

## Discussion

In this large cohort study, we found high rates of ISTH overt DIC score in APL, non-APL AML and ALL patients. We found alterations in levels of several activators, inhibitors of the coagulation and fibrinolytic pathways along with NET markers in patients with acute leukemia compared to healthy controls. EVTF activity was associated with a high risk of bleeding and PAI-1 was associated with a high risk of VTE in acute leukemia patients.

In a previous study, PBMCs (including blast cells) from AML patients had higher levels of TF procoagulant activity compared to PBMCs from ALL patients.[29] In addition, PBMCs from patients with APL had the highest levels of TF procoagulant activity compared to other subtypes of AML. These data are consistent with our findings that EVTF activity levels were highest in APL compared to other types of acute leukemia. In contrast to our results, Sakata et al observed lower PAI-1 activity in APL patients with DIC compared to healthy controls.[45] One possible explanation is that they used an in-house assay to measure PAI-1 activity whereas we used a commercial assay for active PAI-1. The data needs to be confirmed in a larger study. APL cells express annexin A2[66] and S100A10[67] that form a heterotetrameric complex.^54,55^ This complex functions as a receptor for both tPA and plasminogen to effectively generate plasmin in a fibrin-free mechanism on the cell surface.[68] This may explain our results of significantly lower plasma levels of tPA in APL patients compared to healthy controls and the APL and non-APL AML groups. More specifically, the majority of tPA may bind to the annexin A2/S100A10 complexes on APL cells resulting in low plasma levels of tPA in APL patients. This hypothesis needs to be investigated in future studies. Some studies also proposed that APL cells express uPA and uPAR that enhance fibrinolysis independent of the annexin A2/S100A10 pathway[69, 70]. Associations between uPA and uPAR levels and bleeding and VTE need to be investigated in future studies.

We found high levels of cfDNA in acute leukemia patients consistent with previous studies.[29, 47] We also observed low levels of H3Cit-DNA complexes in leukemia patients This is consistent with previous studies demonstrating that granulocytes and neutrophils from acute leukemia patients have reduced capacity to form NETs compared to those from healthy controls.[54–56] It is also notable that APL patients had lower WBC counts that might also affect the levels of H3Cit-DNA complexes in APL patients.

We also found that acute leukemia patients with high EVTF activity had higher WBC count and peripheral blood blast percentage compared to patients with mid to low EVTF activity. These data suggest that WBC and blast cells are the sources of TF+EVs in these patients. Similarly, acute leukemia patients with high EVTF activity had higher D-dimer levels compared to patients with mid to low EVTF activity. This data suggests that EVTF activity is associated with activation of coagulation and fibrinolysis. In contrast, there is no difference of platelet counts between acute leukemia patients with high EVTF activity and mid to low EVTF activity. This data suggests that EVTF activity is not associated with platelet count.

In addition to the known high risk of DIC in APL, we also found high rates of DIC in non-APL AML and ALL patients. While the majority of bleeding and VTE complications occurred early in the clinical course of APL, we found a continued risk of these complications in non-APL AML beyond the first month. We also compared levels of biomarkers between patients with overt DIC and those without DIC. In APL and non-APL AML patients, EVTF activity was significantly higher in patients with overt DIC compared to patients without DIC. There are two possible interpretations of these data. The first is that TF+EVs are the major driver of overt DIC in these patients. The second is that EVTF activity may reflect more severe disease. In APL and non-APL AML patients, WBC count was significantly higher in overt DIC patients compared to patients without DIC. This data suggests that leukocytosis is associated with overt DIC in APL and non-APL AML patients. In addition, peripheral blood blast cell percentage and LDH level were significantly higher in non-APL AML patients with overt DIC compared to patients without DIC. Collectively, these data suggest that leukemia cell burden and disease severity are associated with overt DIC in these patients. Interestingly, levels of cfDNA and PTT were significantly higher in non-APL AML patients with overt DIC compared to patients without DIC. These data suggest that the intrinsic coagulation pathway is possibly activated by cfDNA that results in consumption of the intrinsic coagulation factors in non-APL AML patients with overt DIC leading to prolonged PTT. This needs to be further investigated. Importantly, the incidence of bleeding was significantly higher in non-APL AML patients with overt DIC compared to non-APL AML patients without DIC. We also found lower levels of PS+ EVs in ALL patients with overt DIC compared to ALL patients without DIC. In this regard, it is notable that WBC counts were lower, and platelet counts were significantly lower in ALL patients with overt DIC compared to ALL patients without DIC. Altogether, we believe that the total number of PS+ EVs are low in ALL patients with overt DIC because the sources of PS+ EVs, such as WBC and platelets are low. PAP and D-dimer levels were significantly higher in ALL patients with overt DIC compared to ALL patients without DIC supporting the conclusion that ALL patients with overt DIC have greater activation of fibrinolysis. In addition, we found that hemoglobin levels were significantly lower in ALL patients with overt DIC compared to ALL patients without DIC.

Libourel and colleagues showed that the ISTH overt DIC score and D-dimer significantly predicted venous and arterial thrombosis in non-APL AML patients.[18] In our study, we did not find an association between D-dimer levels and VTE in non-APL AML patients. There are some notable differences between the study by Libourel et al. and our study. First, they included both arterial and venous thrombosis whereas we included only VTE. Second, they excluded catheter-associated VTE whereas we included catheter-associated VTE, as this is a common complication in leukemia patients and is clinically relevant. We believe these differences may have contributed to the different results in the two studies. These observations suggest that different mechanisms of coagulopathy are associated with different types of acute leukemia as well as the types of VTE.

To our knowledge, this is the largest cohort study evaluating the association between different biomarkers of activation and inhibition of the coagulation and fibrinolytic pathways with bleeding and VTE in acute leukemia patients. We found an association between levels of EVTF activity and risk of bleeding, and between PAI-1 levels with VTE in acute leukemia patients. We hypothesize that TF+EVs contribute to over-activation of coagulation that can ultimately result in bleeding due to consumption of coagulation factors in acute leukemia patients. In contrast, PAI-1 inhibits fibrinolysis which may contribute to VTE. Our data suggests that acute leukemia patients may have a sensitive balance between bleeding and VTE that is driven by EVTF activity and PAI-1, respectively. It is also notable that in our study acute leukemia patients showed much higher EVTF activity than levels reported in previous smaller studies.[41, 42] One possible explanation is that different anticoagulants were used to draw blood. In our study, we used EDTA whereas previous studies used sodium citrate.[41, 42]

There are some limitations in the current study. First, there are only 30 healthy control plasma although we have a total of 358 acute leukemia patients’ plasma. The COVID-19 pandemic made it difficult to collect enough number of healthy control plasma. Second, VTE and bleeding are dynamic events in the clinical course of leukemia patients, and this risk is determined by several patient and treatment related factors. Our study did not examine the role of chemotherapy, central venous catheters, and other complications such as infection that might contribute to VTE and/or bleeding risk. We were also not able to capture laboratory parameters at the time of VTE and bleeding. Smaller event rates also preclude examining the associations between biomarkers with outcomes in sub-types of leukemia. Third, 27 out of 29 APL patients received at least one dose of ATRA treatment before sample collection as ATRA is usually administered immediately upon suspicion of APL, before these subjects were enrolled in the study. Previous studies have shown that ATRA reduces TF protein levels in APL cells.[30, 33–36] Therefore, EVTF activity in APL patients might be affected by ATRA treatment in our study. Fourth, the number of no-DIC patients in the APL group were only three and this can skew the results.

In summary, we have shown that APL patients had the highest levels of EVTF activity and PAI-1, and lowest levels of tPA among different types of acute leukemia. ALL patients had the highest levels of cfDNA. Our data suggests that high levels of EVTF activity, WBC, PT and LDH were associated with increased risk of bleeding in acute leukemia patients. High levels of PAI-1 were associated with increased risk of VTE in acute leukemia patients. Future studies investigating these biomarkers in serial plasma samples and the impact of chemotherapy on these biomarkers will help in understanding the mechanisms of coagulopathy in acute leukemia.

## Data Availability

All data produced in the present study are available upon reasonable request to the authors.

## Acknowledgments

This study was supported by the National Heart, Lung, and Blood Institute (N.M. 1R35HL155657) and the John C. Parker Professorship (N.M.). RG received research funding from the National Heart, Lung, and Blood Institute (K08 HL159290) and the American Society of Hematology. We thank Amanda Mullen for assistance with sample banking and retrieval.

## Authorship Contributions

Y.H, N.M. and R.G. designed the study; S.J.A. performed the experiments; K.B, S.D, N.B, L.H helped with data collection, Y.H., S.J.A., Y.C performed statistical analyses, Y.H. and R.G. wrote the article, which was reviewed and edited by all other authors.

