## Supplemental materials for "Biomarkers of bleeding and venous thromboembolism in patients with acute leukemia"

#### Supplementary materials

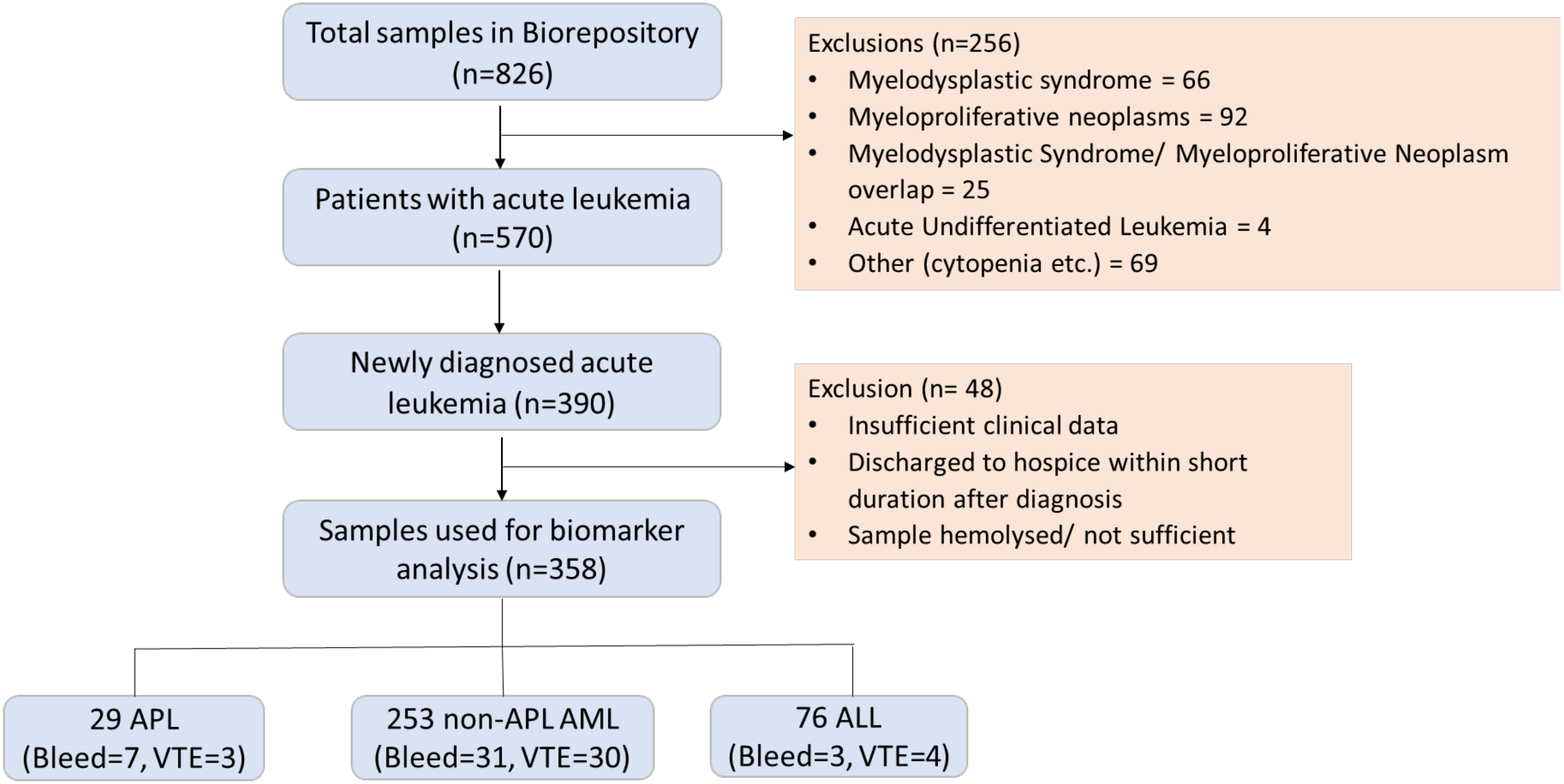


**Supplementary Figure 1. Flow chart to demonstrate inclusion and exclusion of patients in the study.**

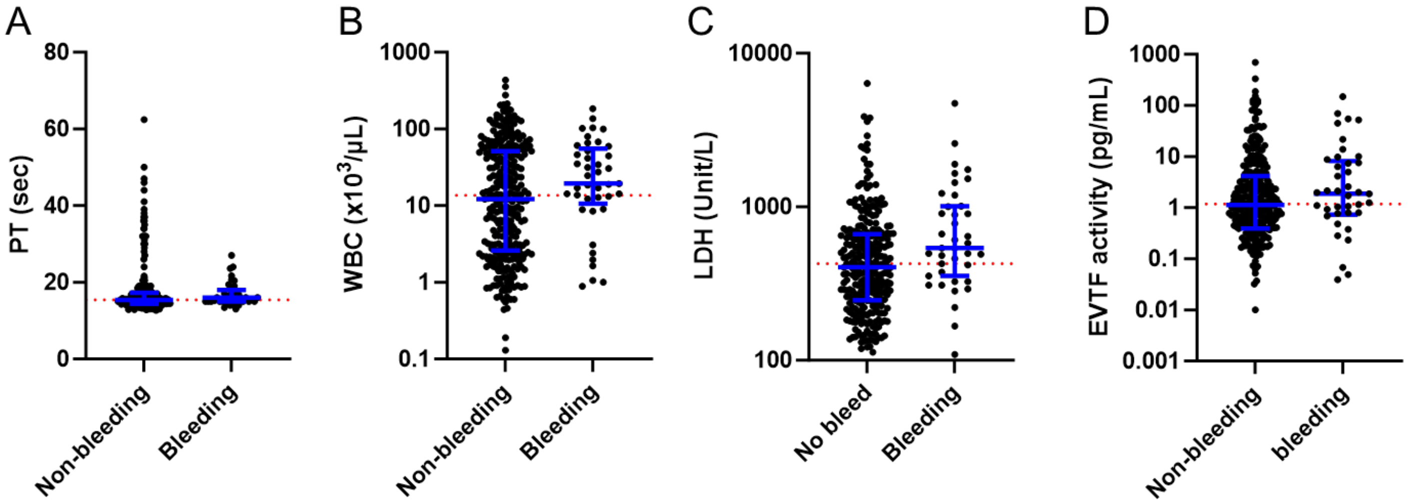


| | A Non-bleeding | A Bleeding | B Non-bleeding | B Bleeding | C No bleed | C Bleeding | D Non-bleeding | D bleeding |
|---|---|---|---|---|---|---|---|---|
| Above median | 48.9% (154/315) | 56.1% (23/41) | 47.6% (151/317) | 68.3% (28/41) | 47.5% (133/280) | 66.7% (26/39) | 49.0% (154/314) | 56.1% (23/41) |

**Supplementary Figure 2. Levels of biomarkers between acute leukemia patients with and without bleeding.** (A) PT, (B) WBC, (C) LDH and (D) EVTF activity levels in acute leukemia patients with and without bleeding are shown. Blue lines indicate median ± interquartile range. Red dotted lines indicate the median level of all patients.

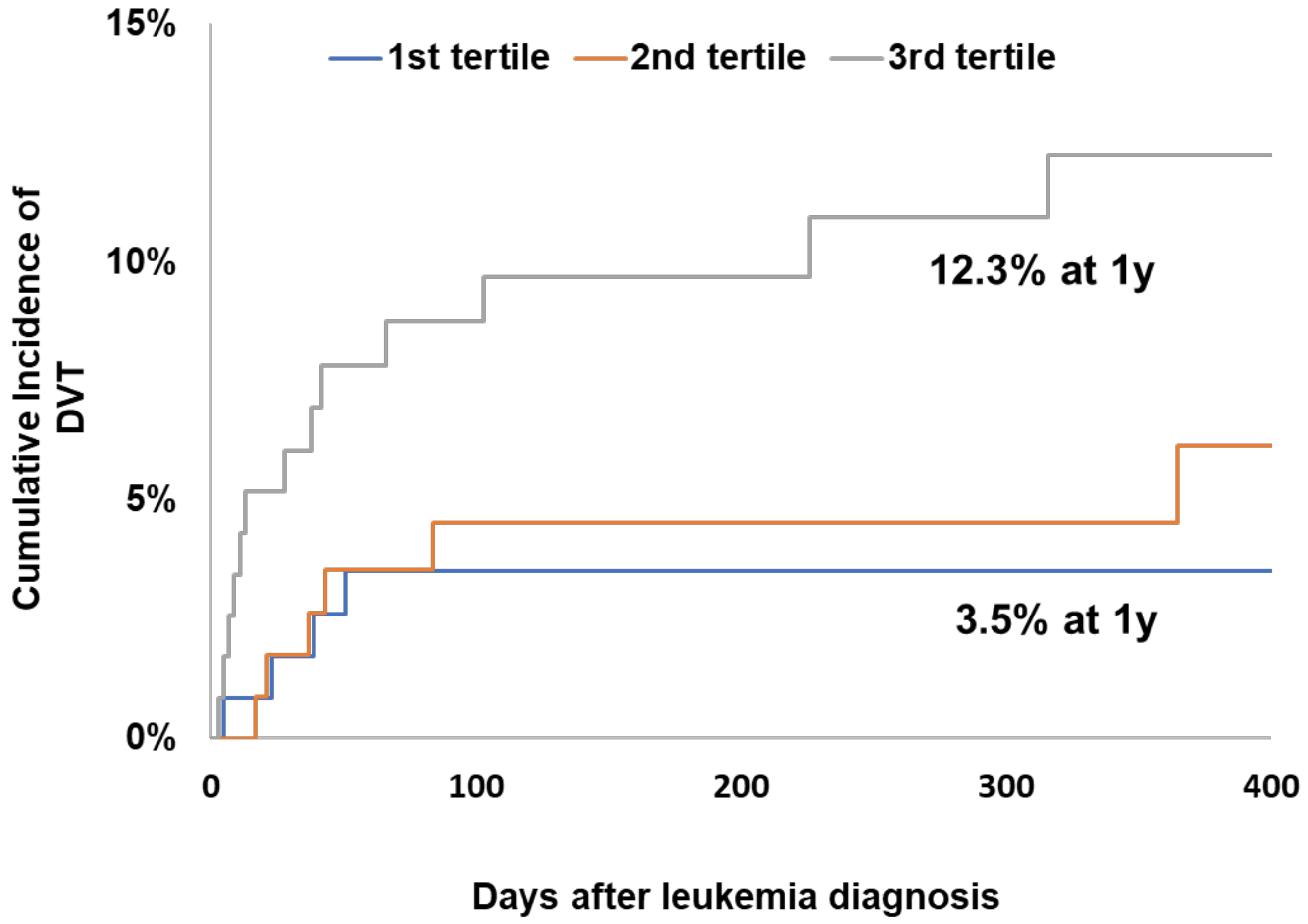


**Supplementary Figure 3. Cumulative incidence of deep vein thrombosis by tertiles of plasminogen activator inhibitor-1 levels.**

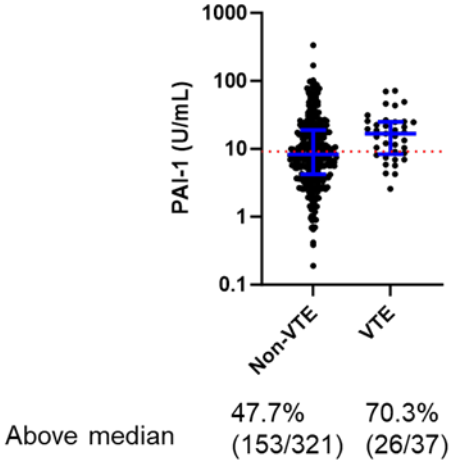


**Supplementary Figure 4. Levels of PAI-1 in acute leukemia patients with and without VTE.** Blue line indicates median ± interquartile range. Red dotted line indicates the median level of all patients.

**Supplementary Table 1. Definitions for disseminated intravascular coagulation, bleeding, and venous thromboembolism outcomes.**

| Outcome | Definition |
|---|---|
| Disseminated intravascular coagulation (DIC) | Defined according to the International Society for Thrombosis and Haemostasis (ISTH) criteria and score ≥5 was defined as overt DIC and score <5 as no-DIC:<br>- Platelet count (≥100 $x10^3$/μL = 0; 50-99 $x10^3$/μL = 1; <50 $x10^3$/μL = 2)<br>- Fibrinogen level (≥100 mg/dL = 0; <100 mg/dL = 1)<br>- Prothrombin time (PT) prolongation above the upper limit of normal (ULN) range (<3 seconds = 0; 3-6 seconds = 1; >6 seconds = 2)<br>- D dimer (<2 times ULN normal [480 ng/mL] = 0; 2-4 times ULN (480-960 ng/mL) = 2; >4 times ULN (960 ng/mL) = 3] |
| Major Bleeding | Defined according to the ISTH classification:<br>- Fatal bleeding<br>- Symptomatic bleeding in critical site/ organ including intracranial, intra-spinal, intraocular, retroperitoneal, intra-articular, pericardial, and intramuscular bleeding with compartment syndrome<br>- Bleeding causing drop in hemoglobin level of ≥2 g/dL or leading to transfusion of ≥2 units of whole blood or red cells |
| Clinically relevant non major bleeding | Defined according to the ISTH classification:<br>- Bleeding requiring medical intervention<br>- Bleeding resulting in hospitalization<br>- Bleeding prompting face-to-face evaluation<br>- Not fulfilling criteria for major bleeding |
| Deep vein thrombosis (DVT) | - Acute symptomatic or incidental lower or upper extremity DVT<br>- Catheter related DVT<br>- Acute symptomatic or incidental visceral vein thrombosis<br>- Acute symptomatic or incidental cerebral vein thrombosis |
| Pulmonary embolism (PE) | - Acute symptomatic PE<br>- Acute incidental PE of the segmental or more proximal pulmonary arterial branches |
| Superficial vein thrombosis | - Acute symptomatic lower or upper extremity thrombosis involving superficial vein |

**Supplementary Table 2. Comparisons between acute leukemia patients with high and mid to low EVTF activity.**

| Parameters | Mid to Low EVTF (n=322) [Median (range)] | High EVTF (n =36) [Median (range)] | P value |
|---|---|---|---|
| WBC count (x $10^3$/µL) | 11,580 [130-435,910] | 59,085 [2,370-171,050] | <0.0001 |
| Peripheral blood blast cell % | 56 [0-97] | 78.3 [37-96] | <0.0001 |
| D-dimer (ng/mL) | 1,138 [135-20,000] | 7,884 [230-20,000] | <0.0001 |
| Platelet count (x $10^3$/µL) | 47,700 [3,400-859,000] | 39,600 [7,100-200,000] | 0.2488 |

**Supplementary Table 3. Comparisons between APL patients with and without overt DIC.**

| Characteristics | No-DIC (n=3) [median (range)] | Overt DIC (n=26) [median (range)] | P value |
|---|---|---|---|
| EVTF activity (pg/mL) | 1.1 (0.7-3.5) | 9.3 (0-188.1) | **0.0353** |
| PS+EVs (nM) | 54.8 (14.4-67.6) | 31.0 (7.5-132.7) | >0.9999 |
| cfDNA (ng/mL) | 985.3 (868-1,103) | 1,144 (799.8-14,766) | NA |
| H3Cit-DNA complex (ng/mL) | 26.9 (0-32.9) | 19.5 (0-424.1) | 0.9087 |
| PAP (ng/mL) | 1,168 (97-15,179) | 6,131 (133-15,349) | 0.8666 |
| tPA (ng/mL) | 0.3 (0-0.9) | 0 (0-3.5) | 0.0999 |
| PAI-1 (U/mL) | 11.0 (7.8-21.2) | 16.7 (2.7-87.7) | 0.5161 |
| D-dimer (ng/mL) | 5,903 (222-6,917) | 16,639 (1,376-20,000) | **0.0112** |
| Fibrinogen (mg/dL) | 289 (281-525) | 167.5 (60-636) | **0.0367** |
| PT (sec) | 14.5 (13.8-15.5) | 17.0 (14.0-27.0) | **0.0435** |
| PTT (sec) | 28 (24-34) | 31 (24-200) | 0.3612 |
| WBC count (x $10^3$/μL) | 0.9 (0.8-0.9) | 3.3 (0.6-79.8) | **0.0085** |
| Peripheral blood blast % | 17 (2.6-77) | 75 (7.5-89) | 0.1550 |
| Platelet count (x $10^3$/μL) | 120 (51.3-131.6) | 29.9 (7.1-112.5) | **0.0011** |
| Creatinine (mg/dL) | 0.6 (0.5-1.7) | 0.9 (0.6-4.4) | 0.3194 |
| Hemoglobin (g/dL) | 9.4 (6.9-10.4) | 8.6 (3.6-12.4) | 0.8283 |
| LDH (Unit/L) | 140 (113-166) | 325 (133-1,898) | NA |
| Bleeding (n, %) | 0 (0) | 7 (26.9) | 0.5575 |
| VTE (n, %) | 0 (0) | 3 (11.5) | >0.9999 |

Abbreviations: cfDNA, cell-free DNA; EVTF, extracellular vesicle tissue factor; H3Cit-DNA, citrullinated histone H3-DNA; LDH, lactate dehydrogenase; NA, not applicable; PAI-1, plasminogen activator inhibitor-1; PAP, plasmin-antiplasmin complex; PS+EVs, phosphatidylserine-positive extracellular vesicles; PT, prothrombin time; PTT, partial thromboplastin time; tPA, tissue plasminogen activator; VTE, venous thromboembolism; WBC, white blood cell

**Supplementary Table 4. Comparisons between non-APL AML patients with and without overt DIC.**

| Characteristics | No-DIC (n=162) [median (range)] | Overt DIC (n=83) [median (range)] | P value |
|---|---|---|---|
| EVTF activity (pg/mL) | 1.1 (0-113) | 2.3 (0-4,086) | **0.0001** |
| PS+EVs (nM) | 56.8 (5.9-939.4) | 53.6 (0-357.2) | 0.2732 |
| cfDNA (ng/mL) | 1,164 (338.7-7,783) | 1,454 (446.2-20,109) | **0.0003** |
| H3Cit-DNA complex (ng/mL) | 32.1 (0-1,552) | 38.2 (0-934) | 0.1901 |
| PAP (ng/mL) | 5,110 (572.7-43,539) | 5,432 (23.6-70,334) | 0.5631 |
| tPA (ng/mL) | 1.8 (0-38.8) | 2.0 (0-19.6) | 0.6449 |
| PAI-1 (U/mL) | 8.0 (0.2-101.9) | 8.9 (0.7-335.4) | 0.1146 |
| D-dimer (ng/mL) | 732 (135-20,000) | 2,182 (624-20,000) | **<0.0001** |
| Fibrinogen (mg/dL) | 456 (181-1,101) | 455 (60-795) | 0.4014 |
| PT (sec) | 15.0 (12.9-47.0) | 16.9 (14.0-62.4) | **<0.0001** |
| PTT (sec) | 33 (19-88) | 36 (21-61) | **0.0002** |
| WBC count (x $10^3$/μL) | 10.3 (0.4-277.3) | 42.0 (0.6-435.9) | **<0.0001** |
| Peripheral blood blast cell (%) | 52 (0-95) | 72 (2.1-96) | **0.0012** |
| Platelet count (x $10^3$/μL) | 66.4 (3.4-862.4) | 30.5 (5.7-132.8) | **<0.0001** |
| Creatinine (mg/dL) | 0.9 (0.4-3.2) | 1.1 (0.5-3.0) | **0.0343** |
| Hemoglobin (g/dL) | 9.0 (2.9-15.1) | 8.7 (3.4-12.8) | 0.3687 |
| LDH (Unit/L) | 360 (109-3,594) | 593.5 (131-5,469) | **<0.0001** |
| Bleeding (n, %) | 14 (8.6) | 15 (18.1) | **0.0373** |
| VTE (n, %) | 17 (10.5) | 12 (14.5) | 0.4054 |

Abbreviations: cfDNA, cell-free DNA; EVTF, extracellular vesicle tissue factor; H3Cit-DNA, citrullinated histone H3-DNA; LDH, lactate dehydrogenase; PAI-1, plasminogen activator inhibitor-1; PAP, plasmin-antiplasmin complex; PS+EVs, phosphatidylserine-positive extracellular vesicles; PT, prothrombin time; PTT, partial thromboplastin time; tPA, tissue plasminogen activator; VTE, venous thromboembolism; WBC, white blood cell

**Supplementary Table 5. Comparisons between ALL patients with and without overt DIC.**

| Characteristics | No-DIC (n=42) [median (range)] | Overt DIC (n=34) [median (range)] | P value |
|---|---|---|---|
| EVTF activity (pg/mL) | 0.7 (0-37.5) | 0.5 (0-52.4) | 0.1064 |
| PS+EVs (nM) | 57.7 (2.7-123.2) | 36.3 (4.5-132.4) | **0.0476** |
| cfDNA (ng/mL) | 1,593 (767.4-22,157) | 1,677 (116.4-29,325) | 0.8407 |
| H3Cit-DNA complex (ng/mL) | 18.9 (0-240.1) | 19.7 (0-1,607) | 0.7649 |
| PAP (ng/mL) | 3,563 (125.4-60,381) | 7677 (561.5-115,720) | **0.0007** |
| tPA (ng/mL) | 0.5 (0-7.5) | 0.5 (0-2.4) | 0.3581 |
| PAI-1 (U/mL) | 10.0 (1.4-57.7) | 8.8 (0.4-69.6) | 0.3103 |
| D-dimer (ng/mL) | 1,045 (233-11,918) | 1,970 (566-16,414) | **0.0024** |
| Fibrinogen (mg/dL) | 401.5 (170.0-856.0) | 374.0 (184.0-687.0) | 0.5694 |
| PT (sec) | 14.4 (12.7-19.2) | 15.0 (12.8-21.6) | 0.0963 |
| PTT (sec) | 32 (24-40) | 33 (25-43) | 0.2146 |
| WBC count (x $10^3$/μL) | 18.3 (0.7-204.6) | 11.7 (0.1-357.0) | 0.3425 |
| Peripheral blood blast cell (%) | 60.5 (1.6-94) | 64 (2.1-97) | 0.8666 |
| Platelet (x $10^3$/μL) | 80.0 (12.6-377.3) | 23.8 (7.9-97.9) | **<0.0001** |
| Creatinine (mg/dL) | 0.9 (0.4-2.1) | 0.9 (0.4-2.2) | 0.7363 |
| Hemoglobin (g/dL) | 9.5 (6.6-17.9) | 8.1 (4.6-11.7) | **0.0002** |
| LDH (Unit/L) | 481 (119-2,563) | 476.5 (123-6,374) | 0.6532 |
| Bleeding (n, %) | 0 (0) | 3 (8.8) | 0.0851 |
| VTE (n, %) | 3 (7.1) | 1 (2.9) | 0.6235 |

Abbreviations: cfDNA, cell-free DNA; EVTF, extracellular vesicle tissue factor; H3Cit-DNA, citrullinated histone H3-DNA; LDH, lactate dehydrogenase; PAI-1, plasminogen activator inhibitor-1; PAP, plasmin-antiplasmin complex; PS+EVs, phosphatidylserine-positive extracellular vesicles; PT, prothrombin time; PTT, partial thromboplastin time; tPA, tissue plasminogen activator; VTE, venous thromboembolism; WBC, white blood cell

**Supplementary Table 6. Characteristics of acute leukemia patients with and without bleeding.**

| Variable | No Bleeding (n=317) | Bleeding (n=41) | P value |
|---|---|---|---|
| **Age at Diagnosis [median, (range)]** | 59 (19-89) | 57 (30-75) | 0.841 |
| **Sex (n, %)** | | | |
| Male | 175 (56.1) | 28 (60.9) | 0.541 |
| **Race (n, %)** | | | |
| Non-Hispanic White | 232 (74.4) | 32 (69.6) | 0.502 |
| Black | 70 (22.4) | 11 (23.9) | |
| Other/Hispanic or Latino | 10 (3.2) | 3 (6.5) | |
| **Body mass index (n, %)** | 31 (7.4) | 30.1 (8.1) | 0.295 |
| **Parameters [median (range)]** | | | |
| EVTF activity (pg/mL) | 1.1 (0-695.6) | 1.9 (0-149.5) | 0.111 |
| PS+EVs (nM) | 54.6 (0-939.4) | 49.9 (4.5-357.2) | 0.375 |
| cfDNA (ng/mL) | 1,226 (116-29,325) | 1,341 (463-14,766) | 0.124 |
| H3Cit-DNA complex (ng/mL) | 50(0-934) | 22 (0-436) | 0.141 |
| PAP (ng/mL) | 5,014 (24-115,720) | 6,251 (1,181-43,539) | 0.091 |
| tPA (ng/mL) | 0.7 (0-38.8) | 1.4 (0-12.6) | 0.476 |
| PAI-1 (U/mL) | 9.1 (0.4-335.4) | 9.5 (0.2-87.7) | 0.208 |
| D-dimer (ng/mL) | 1,288 (135-20,000) | 2,182 (212-20,000) | **0.039** |
| Fibrinogen (mg/dL) | 421 (60-1,101) | 441 (60-722) | 0.382 |
| PT (sec) | 15.1 (12.7-62.4) | 16.0 (13.0-27.0) | **0.027** |
| PTT (sec) | 33 (19-200) | 33 (24-51) | 0.807 |
| WBC count (x $10^3$/µL) | 12.1 (0.1-435.9) | 19.0 (0.9-136.0) | 0.148 |
| Peripheral blood blast cell (%) | 60 (0-97) | 70 (0-91) | 0.477 |
| Platelet (x $10^3$/µL) | 47.7 (3.4-862.4) | 35.0 (11.1-285.2) | 0.114 |
| Creatinine (mg/dL) | 0.9 (0.4-3.2) | 0.8 (0.4-4.4) | 0.977 |
| Hemoglobin (g/dL) | 8.7 (2.9-17.9) | 8.5 (3.8-13.6) | 0.111 |
| LDH (Unit/L) | 404 (110-6,374) | 539 (109-4,714) | **0.006** |
| **History of Bleeding (n, %)** | | | |
| Yes | 4 (1.3%) | 1 (2.2%) | 1.261 |

**Supplementary Table 6. Characteristics of acute leukemia patients with and without bleeding. (continued)**

| Variable | No Bleeding (n=317) | Bleeding (n=41) | P value |
|---|---|---|---|
| **Comorbidities (n, %)** | | | |
| None | 114 (36.5) | 19 (41.3) | 0.523 |
| 1 | 78 (25) | 8 (17.4) | |
| ≥2 | 120 (38.5) | 19 (41.3) | |

Abbreviations: cfDNA, cell-free DNA; EVTF, extracellular vesicle tissue factor; H3Cit-DNA, citrullinated histone H3-DNA; LDH, lactate dehydrogenase; PAI-1, plasminogen activator inhibitor-1; PAP, plasmin-antiplasmin complex; PS+EVs, phosphatidylserine-positive extracellular vesicles; PT, prothrombin time; PTT, partial thromboplastin time; tPA, tissue plasminogen activator; WBC, white blood cell

**Supplementary Table 7. Characteristics of acute leukemia patients with and without venous thromboembolism.**

| Variable | No VTE (n=321) | VTE (n=37) | P value |
|---|---|---|---|
| **Age at Diagnosis [median (range)** | 59 (19-89) | 54 (20-72) | 0.135 |
| **Sex (n, %)** | | | |
| Male | 185 (58.2) | 18 (45.0) | 0.113 |
| **Race (n, %)** | | | |
| Non-Hispanic White | 233 (73.3) | 31 (77.5) | 0.423 |
| Black | 72 (22.6) | 9 (22.5) | |
| Other/Hispanic or Latino | 13 (4.1) | 0 (0) | |
| **Body mass index (n, %)** | 30.9 (7.6) | 31.3 (6.3) | 0.426 |
| **Parameters [median (range)]** | | | |
| EVTF activity (pg/mL) | 1.1 (0-695.6) | 1.8 (0-334.2) | 0.156 |
| PS+EVs (nM) | 53.4 (0-939.4) | 60.1 (7.7-326) | 0.346 |
| cfDNA (ng/mL) | 1,263 (116-29,325) | 1,056 (430-6,114) | 0.111 |
| H3Cit-DNA complex (ng/mL) | 29 (0-1607) | 27 (0-404) | 0.419 |
| PAP (ng/mL) | 5,445 (24-115,720) | 4,288 (133-83,099) | 0.123 |
| tPA (ng/mL) | 0.8 (0-38.8) | 0.2 (0-12.6) | 0.067 |
| PAI-1 (U/mL) | 8.3 (0.2-335.4) | 16.7 (2.6-71.6) | **0.002** |
| D-dimer (ng/mL) | 1,383 (135-20,000) | 1,154 (233-20,000) | 0.943 |
| Fibrinogen (mg/dL) | 427 (60-1,100) | 378 (98-882) | 0.192 |
| PT (sec) | 15.2 (12.7-62.4) | 15.0 (13.6-24.6) | 0.43 |
| PTT (sec) | 33 (19-200) | 32 (24-50) | 0.975 |
| WBC count (x $10^3$/μL) | 13.2 (0.1-358.1) | 18.3 (0.9-435.9) | 0.168 |
| Peripheral blood blast cell (%) | 60 (0-97) | 57 (6-96) | 0.884 |
| Platelet (x $10^3$/μL) | 46 (3-862) | 41 (10-353) | 0.695 |
| Creatinine (mg/dL) | 0.9 (0.4-4.4) | 0.9 (0.5-1.8) | 0.589 |
| Hemoglobin (g/L) | 8.6 (2.9-17.9) | 9.0 (3.6-12.8) | 0.692 |
| LDH (Unit/L) | 424 (109-6,374) | 401 (110-5,469) | 0.965 |
| **History of VTE (n, %)** | | | |
| Yes | 14 (4.4%) | 3 (7.5%) | 0.385 |

**Supplementary Table 7. Characteristics of acute leukemia patients with and without venous thromboembolism. (continued)**

| Variable | No VTE (n=321) | VTE (n=37) | P value |
|---|---|---|---|
| **Comorbidities (n, %)** | | | |
| None | 117 (36.8) | 16 (40.0) | 0.867 |
| 1 | 76 (23.9) | 10 (25.0) | |
| ≥2 | 125 (39.3) | 14 (35.0) | |

Abbreviations: cfDNA, cell-free DNA; EVTF, extracellular vesicle tissue factor; H3Cit-DNA, citrullinated histone H3-DNA; LDH, lactate dehydrogenase; PAI-1, plasminogen activator inhibitor-1; PAP, plasmin-antiplasmin complex; PS+EVs, phosphatidylserine-positive extracellular vesicles; PT, prothrombin time; PTT, partial thromboplastin time; tPA, tissue plasminogen activator; VTE, venous thromboembolism; WBC, white blood cell

**Supplementary Table 8. Biomarkers associated with bleeding and VTE risk in non-APL AML.**

| Biomarker | Bleeding# | | VTE* | |
|---|---|---|---|---|
| | HR (95% CI) | p-value | HR (95% CI) | p-value |
| EVTF activity | **2.35 (0.98 - 5.66)** | **0.057** | 1.27 (0.58 - 2.77) | 0.546 |
| PS + EVs | 1.25 (0.61 - 2.56) | 0.534 | 0.89 (0.43 - 1.85) | 0.758 |
| cfDNA | 1.99 (0.97 - 4.07) | 0.061 | 0.55 (0.28 - 1.08) | 0.081 |
| H3Cit-DNA complexes | 0.76 (0.40 - 1.46) | 0.414 | 0.54 (0.27 - 1.09) | 0.084 |
| PAP | 1.30 (0.64 - 2.62) | 0.470 | 0.84 (0.41 - 1.70) | 0.623 |
| tPA | 0.88 (0.42 - 1.82) | 0.726 | **0.47 (0.24 - 0.93)** | **0.029** |
| PAI-1 | 1.32 (0.65 - 2.66) | 0.444 | **4.18 (1.47 - 11.87)** | **0.007** |
| D-dimer | 1.32 (0.65 - 2.68) | 0.435 | 1.01 (0.50 - 2.06) | 0.979 |
| Fibrinogen | 0.66 (0.34 - 1.28) | 0.217 | 0.74 (0.37 - 1.49) | 0.402 |
| PT | 1.87 (0.85 - 4.13) | 0.120 | 1.55 (0.67 - 3.56) | 0.306 |
| PTT | 0.66 (0.34 - 1.29) | 0.226 | 1.35 (0.61 - 3.00) | 0.454 |
| WBC count | **2.42 (1.04 - 5.62)** | **0.041** | 0.94 (0.46 - 1.89) | 0.857 |
| Peripheral blood blast cell | 1.08 (0.54 - 2.17) | 0.827 | 0.97 (0.45 - 2.08) | 0.928 |
| Platelet count | 0.56 (0.29 - 1.09) | 0.086 | 0.75 (0.37 - 1.50) | 0.411 |
| Creatinine | 1.14 (0.49 - 2.63) | 0.762 | 1.10 (0.50 - 2.43) | 0.805 |
| Hemoglobin | 0.72 (0.38 - 1.39) | 0.330 | 1.44 (0.67 - 3.10) | 0.353 |
| LDH | **3.50 (1.34 - 9.17)** | **0.011** | 1.08 (0.52 - 2.22) | 0.842 |

### Multivariable model was adjusted for age, sex, race/ethnicity,

*Multivariable model was adjusted for age, sex, race/ethnicity, body mass index, history of venous thromboembolism and comorbidities.

Abbreviations: cfDNA, cell-free DNA; EVTF, extracellular vesicle tissue factor; H3Cit-DNA, citrullinated histone H3-DNA; LDH, lactate dehydrogenase; PAI-1, plasminogen activator inhibitor-1; PAP, plasmin-antiplasmin complex; PS+EVs, phosphatidylserine-positive extracellular vesicles; PT, prothrombin time; PTT, partial thromboplastin time; tPA, tissue plasminogen activator; WBC, white blood cell; 95% CI, 95% confidence interval

**Supplementary Table 9. Biomarkers associated with intracranial bleeding in patients with acute leukemia**

| Biomarker | Comparison | Univariable | |
|---|---|---|---|
| | | HR (95% CI) | p-value |
| EVTF activity | Tertile 2 and 3 vs. 1 | 6.16 ( 0.82 - 46.34) | **0.0774** |
| PS + EVs | Tertile 2 and 3 vs. 1 | 0.82 ( 0.26 - 2.54) | 0.7259 |
| cfDNA | Tertile 2 and 3 vs. 1 | 2.62 ( 0.58 - 11.88) | 0.2114 |
| H3Cit-DNA complexes | Tertile 2 and 3 vs. 1 | 1.11 ( 0.33 - 3.75) | 0.8670 |
| PAP | Tertile 2 and 3 vs. 1 | 0.93 ( 0.28 - 3.06) | 0.9063 |
| tPA | Tertile 2 and 3 vs. 1 | 0.29 ( 0.09 - 0.90) | **0.0318** |
| PAI-1 | Tertile 2 and 3 vs. 1 | 0.73 ( 0.24 - 2.28) | 0.5930 |
| D-dimer | Tertile 2 and 3 vs. 1 | 2.56 ( 0.58 - 11.40) | 0.2171 |
| Fibrinogen | Tertile 2 and 3 vs. 1 | 0.27 ( 0.09 - 0.84) | **0.0234** |
| PT | Tertile 2 and 3 vs. 1 | 1.67 ( 0.37 - 7.42) | 0.5023 |
| PTT | Tertile 2 and 3 vs. 1 | 0.81 ( 0.25 - 2.69) | 0.7357 |
| WBC count | Tertile 2 and 3 vs. 1 | 1.92 ( 0.43 - 8.64) | 0.3931 |
| Peripheral blood blast cell | Tertile 2 and 3 vs. 1 | 3.90 ( 0.51 - 29.97) | 0.1912 |
| Platelet count | Tertile 2 and 3 vs. 1 | 0.85 ( 0.27 - 2.63) | 0.7729 |
| Creatinine | Tertile 2 and 3 vs. 1 | 3.45 ( 0.45 - 26.48) | 0.2330 |
| Hemoglobin | Tertile 2 and 3 vs. 1 | 0.55 ( 0.18 - 1.67) | 0.2875 |
| LDH | Tertile 2 and 3 vs. 1 | 2.14 ( 0.48 - 9.61) | 0.3212 |

Abbreviations: cfDNA, cell-free DNA; EVTF, extracellular vesicle tissue factor; H3Cit-DNA, citrullinated histone H3-DNA; LDH, lactate dehydrogenase; PAI-1, plasminogen activator inhibitor-1; PAP, plasmin-antiplasmin complex; PS+EVs, phosphatidylserine-positive extracellular vesicles; PT, prothrombin time; PTT, partial thromboplastin time; tPA, tissue plasminogen activator; WBC, white blood cell; 95% CI, 95% confidence interval
